# Effect of dialysis modality on the survival of end-stage renal disease patients starting dialysis in Sabah from 2007 to 2017: a retrospective cohort study

**DOI:** 10.1101/2021.07.26.21261164

**Authors:** Thamron Keowmani, Anis Kausar Ghazali, Najib Majdi Yaacob, Koh Wei Wong

## Abstract

**Background:** The effect of dialysis modality on the survival of end-stage renal disease patients is a major public health interest.

**Methods:** In this retrospective cohort study, all adult end-stage renal disease patients receiving dialysis treatment in Sabah between January 1, 2007 and December 31, 2017 as identified from the Malaysian Dialysis and Transplant Registry were evaluated and followed up through December 31, 2018. The endpoint was all-cause mortality. The observation time was defined as the time from the date of dialysis initiation after the onset of end-stage renal disease to whichever of the following that came first: date of death, date of transplantation, date of last follow-up, date of recovered kidney function, or December 31, 2018. Weighted Cox regression was used to estimate the effect of dialysis modality. Analyses were restricted to patients with complete data on all variables.

**Results:** 1,837 patients began hemodialysis and 156 patients started with peritoneal dialysis, yielding 7,548.10 (potential median 5.48 years/person) and 747.98 (potential median 5.68 years/person) person-years of observation. 3.1% of patients were lost to follow-up. The median survival time was 5.8 years (95% confidence interval: 5.4, 6.3) among patients who started on hemodialysis and 7.0 years (95% confidence interval: 5.9, indeterminate) among those who started on peritoneal dialysis. The effect of dialysis modality was not significant after controlling for confounders. The average hazard ratio was 0.80 (95% confidence interval: 0.61, 1.05) with hemodialysis as a reference.

**Conclusion:** There was no evidence of a difference in mortality between hemodialysis and peritoneal dialysis.

## Introduction

End-stage renal disease (ESRD), also known as renal failure, occurs when chronic kidney disease (CKD), the gradual loss of kidney function, reaches an advanced stage. ESRD is a growing public health problem globally. The quantification of the ESRD burden is largely based on national registries of ESRD patients who are receiving renal replacement therapy (RRT). This group of patients represents the treated ESRD population [1].

RRT is a life-saving treatment for patients with ESRD. It is provided by kidney transplantation or long-term dialysis. It is known that transplantation extends lives, provides better quality of life, and is more cost-effective than dialysis. However, there is an insufficient number of kidney donors, especially in countries with relatively high incidence rates of treated ESRD. Therefore, most patients require chronic dialysis [2].

Dialysis can be provided by hemodialysis (HD) or peritoneal dialysis (PD). The latter is considered more cost-effective than the former but used less often. One possible explanation is the larger number of HD centres available. An important question is whether a survival advantage exists between HD and PD. Demonstrating a mortality benefit or lack of benefit would affect dialysis choice and clinical management policies prior to ESRD development.

Numerous studies have been conducted to compare the survival outcomes between HD and PD, but there is yet to be any conclusive evidence to favor one over the other. Most findings have come from retrospective cohort studies based on national health or insurance databases [3,4]. There was only one notable clinical trial comparing the survival between HD and PD in ESRD patients, of which PD was reported to lead to better survival [5].

Studies comparing survival between HD and PD patients mostly come from the Western world [3,4,6,7] and East Asian countries [8-10]. There is only one similar study that was conducted in Southeast Asia, which was performed in Singapore [11]. The difference in socioeconomic status, culture, race, access to treatment, and other nonmodifiable factors between the different populations studied make generalization across populations difficult. As such, survival studies specific to ESRD patients in Sabah are needed.

Sabah is one of the 13 states of Malaysia. It is situated on the northern portion of Borneo Island, which is separated from peninsular Malaysia by the South China Sea. The racial composition between the two parts of the country is vastly different. Economically, Sabah is among the poorest state in the country. The dialysis treatment rates in the state over the years were also among the lowest, ranging from 68 per million population (PMP) in 2007 to 125 PMP in 2016 [12]. To date, there are no known published studies on survival in this population.

The first objective of this study was to compare the survival of ESRD patients receiving dialysis in Sabah who were started on HD with that of those who were started on PD. Based on existing evidence, it was hypothesized that there would be no difference in survival between the two modalities after controlling for confounders. The second objective was to identify the factors associated with all-cause mortality.

## Methods

### Study design and setting

This is a retrospective cohort study of new ESRD patients who received dialysis treatment in Sabah between January 1, 2007 and December 31, 2017. Incident dialysis patients in Sabah from 2007 through 2017 were evaluated and followed up through December 31, 2018. The data used in this study were obtained from the Malaysian Dialysis and Transplant Registry (MDTR). A total of 49 participating clinical sites in Sabah contributed the data to MDTR during the observation period.

Established in 1993, the MDTR collects information on patients with ESRD on RRT in Malaysia and the objective of the MDTR is to evaluate the health outcomes of such patients. The patients were followed from the time they were registered until death. The outcome of death was ascertained for all patients registered, including those who were lost to follow-up, via the National Registration Department (NRD). In accordance with the Private Healthcare Facilities Act 1998 (AKTA 586), all dialysis health facilities are required to submit data to the MDTR. In Sabah, only those patients who have been on dialysis for more than 3 months were reported.

### Eligibility criteria

All ESRD patients aged 18 years or older who started either HD or PD in Sabah as identified from MDTR between January 1, 2007, and December 31, 2017 were eligible for the study. There was no sampling. All patients who fulfilled the above eligibility criteria were included and formed the study cohort. The cohort was then “followed” until December 31, 2018.

### Variables

The outcome of interest was survival time, and the endpoint was death from any cause. The observation time was defined as the time from the date of dialysis initiation after the onset of ESRD to whichever of the following came first: date of death, date of transplantation, date of last follow-up only if there was no date of death reported, date of recovered kidney function, or December 31, 2018. A switch in dialysis modality was not considered a censoring event to mimic the intention-to-treat method of analysis.

The exposure of interest was dialysis modality. The other predictors studied were age, sex, body mass index (BMI), diabetes mellitus (DM) comorbidity, cardiovascular disease (CVD) comorbidity (including ischemic heart disease (IHD), congestive cardiac failure (CCF), and other cardiac comorbidities), serum creatinine, serum potassium, serum albumin, serum calcium, serum phosphate, serum alkaline phosphatase (ALP), total cholesterol, and hemoglobin (Hb). All laboratory values were longitudinal data measured after dialysis initiation. Their means were used for analyses.

### Pre-study sample size calculation

Ultimately, the number of patients who met the eligibility criteria determined the sample size. However, prestudy sample size calculation for the first objective was also performed using `power cox` command in Stata/SE 15.1 (StataCorp, College Station, TX, USA). Given an overall probability of death of 13.9%, a prevalence of PD of approximately 10% [12] and that, all confounders explained 25% of the dialysis modality selection [13], it was estimated that at least 1,742 subjects were needed to ensure a power of 80% to detect an alternative Ha: *β*1 = -0.6931 (corresponding to HR of 0.5) using a two-sided test with a 0.05 level of significance.

### Statistical analyses

All analyses were restricted to patients with complete data for all variables predetermined for regression analyses. This may be a source of bias. To show that the complete cases are typical of the whole sample, demographic characteristics and laboratory values between them were compared.

For descriptive purposes, the number of patients who died and total person-years at risk were tabulated according to dialysis modality. The naïve median follow-up time was calculated by using all the observation times and determining the median. The disadvantage of this method is that a study with a long observation period, but many early deaths would appear not to have a long follow-up time. The preferred alternative is “potential” median follow-up time.

“Potential” median follow-up time was calculated by using the “Reverse” Kaplan-Meier (KM) method. First, the censoring indicators were switched. Then, the “survival” estimate was computed using the reversed censoring indicators, and the median “survival” was found [14].

For the first objective, the characteristics of patients who were started on HD were compared to those who were started on PD. Welch’s two-sample *t*-test and Cohen’s *d* were used to compare continuous covariates and the chi-square test of independence was used to compare categorical covariates. The effect of PD versus that of HD was assessed, adjusted for all the other predictors, by using multivariable weighted Cox regression (WCR). There was no variable selection.

For the second objective, multivariable WCR was fitted after a model-building process. The model building process consisted of five steps: first, univariable Cox proportional hazards (PH) were fitted for each predictor; second, variable selection was performed by the backward stepwise AIC method (*p*-value < 0.05 was not a major determinant for variable selection); third, selected interaction terms were tested (`Modality`-`age`, `Modality`-`comorbidDM`, `Modality`-`comorbidCVD`); fourth, model adequacy was assessed; and fifth, model specification was modified.

WCR is a parsimonious option to account for time-dependent effects. It yields an estimate of the average hazard ratio (AHR) for each covariate. This can be interpreted as an average effect of that covariate over the observation period. It permits a clear-cut answer to the question of which treatment or level of a prognostic factor is favorable, independent of the assumption of proportional hazards. A robust (Lin-Wei) covariance matrix was used for significance testing in all WCRs [15,16].

Every regression analysis was started with the Cox PH model. WCR was used in place of the model only after the assessment of model adequacy. Both univariable and multivariable Cox PH models were tested for the PH assumption. In addition, every fitted multivariable Cox PH model was checked for the following: linearity of each continuous covariate against the log relative hazard, influential outliers, and overall goodness of fit. Each continuous covariate was standardized, for ease of interpretation, by first centering its values around the mean and then dividing by its standard deviation. Each standardized X-coefficient was then interpreted as the change in hazard or death rate for every one-SD change in X, when all other variables were held constant.

Linearity was checked by plotting Martingale residuals from the fitted model against the covariate, overlaid by loess smooth curve with the 95 percent confidence interval (CI). Slight deviation from linearity (a straight line) was permitted. Influential outliers were identified by plotting DFBETASs against subjects’ identification numbers. Any values > |1| were considered influential. Overall goodness-of-fit was assessed by plotting the cumulative hazards (derived from the KM estimator using Cox-Snell residuals in place of observed survival time) against Cox-Snell residuals. The degree of superimposition determined the goodness-of-fit. Proportional hazards assumption was assessed by tests based on Schoenfeld residuals, which tested for slope of *β*(*t*) = 0.

All analyses were performed by using R version 3.6.2 (R Core Team, VA, Austria).

## Results

Of the 3,628 eligible patients, 1,993 were complete cases. Figure 1 shows the process by which patients were included in the study. The characteristics of the complete cases were typical of the characteristics of all cases as shown in Table 1 and Table 2. Most of the patients were aged between 40 and 68 years old, males (56.7%), having at most secondary education (49.5%), and having monthly family income less than RM 1,000 (48.4%). Table 3 shows the number of deaths and total person-years at risk by dialysis modality for the complete cases. During the 8,296.7 person-years (potential median 5.5 years/person) of follow-up, 893 (44.8%) patients died, 61 (3.1%) were lost to follow-up, 17 (0.9%) received transplant, and 5 (0.3%) recovered. The naïve median follow-up time was 3.5 years.

**Figure 1.**
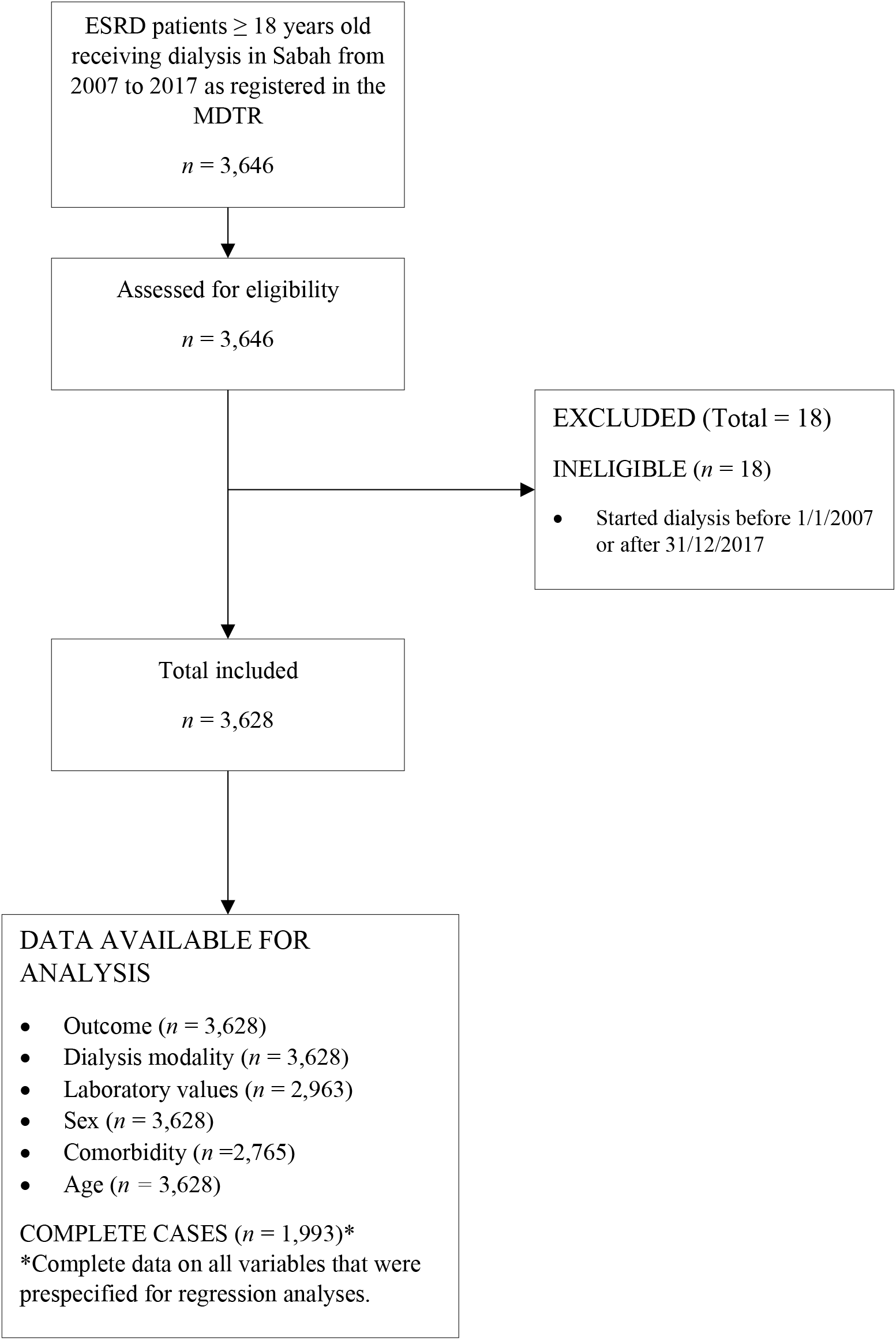
Flow diagram of patients included in the study.

**Table 1.** Demographic characteristics of patients.

|  | All cases |  |  | Complete cases<sup>a</sup> |  |  |
| --- | --- | --- | --- | --- | --- | --- |
|  | *n* | *n* (%) | Mean (SD) | *n* | *n* (%) | Mean (SD) |
| Age (years) | 3628 |  | 53.39<br>(13.92) | 1993 |  | 53.79<br>(13.50) |
| Sex | 3628 |  |  | 1993 |  |  |
| Male |  | 2055<br>(56.6) |  |  | 1130<br>(56.7) |  |
| Female |  | 1573<br>(43.4) |  |  | 863<br>(43.3) |  |
| BMI (kg/m<sup>2</sup>) | 2832 |  | 24.64<br>(5.10) | 1993 |  | 24.75<br>(4.67) |
| DM Comorbidity | 2765 |  |  | 1993 |  |  |
| Yes |  | 1439<br>(52.0) |  |  | 1034<br>(51.9) |  |
| No |  | 1326<br>(48.0) |  |  | 959<br>(48.1) |  |
| CVD Comorbidity | 2765 |  |  | 1993 |  |  |
| Yes |  | 224<br>(8.1) |  |  | 152<br>(7.6) |  |
| No |  | 2541<br>(91.9) |  |  | 1841<br>(92.4) |  |
| Initial dialysis modality | 3628 |  |  | 1993 |  |  |
| HD |  | 3212<br>(88.5) |  |  | 1837<br>(92.2) |  |
| PD |  | 416<br>(11.5) |  |  | 156<br>(7.8) |  |

<sup>a</sup> Complete data for all variables prespecified for regression analyses.

**Table 2.** Mean laboratory values after dialysis initiation.

|  | All cases (*n* = 2963) | Complete cases (*n* = 1993) |
| --- | --- | --- |
|  | Mean (SD) | Mean (SD) |
| Serum Creatinine (µm/L) | 823.71 (301.84) | 812.31 (288.64) |
| Serum Albumin (g/L) | 36.85 (6.84) | 37.21 (6.51) |
| Serum Potassium (mmol/L) | 4.45 (0.86) | 4.49 (0.84) |
| Serum Calcium (mmol/L) | 2.17 (0.25) | 2.18 (0.24) |
| Serum Phosphate (mmol/L) | 1.78 (0.57) | 1.78 (0.54) |
| Serum ALP (U/L) | 112.24 (75.94) | 113.20 (79.53) |
| Hb (g/dL) | 9.81 (1.68) | 9.85 (1.67) |
| Total Cholesterol (mmol/L) | 4.46 (1.85) | 4.40 (1.13) |

**Table 3.**
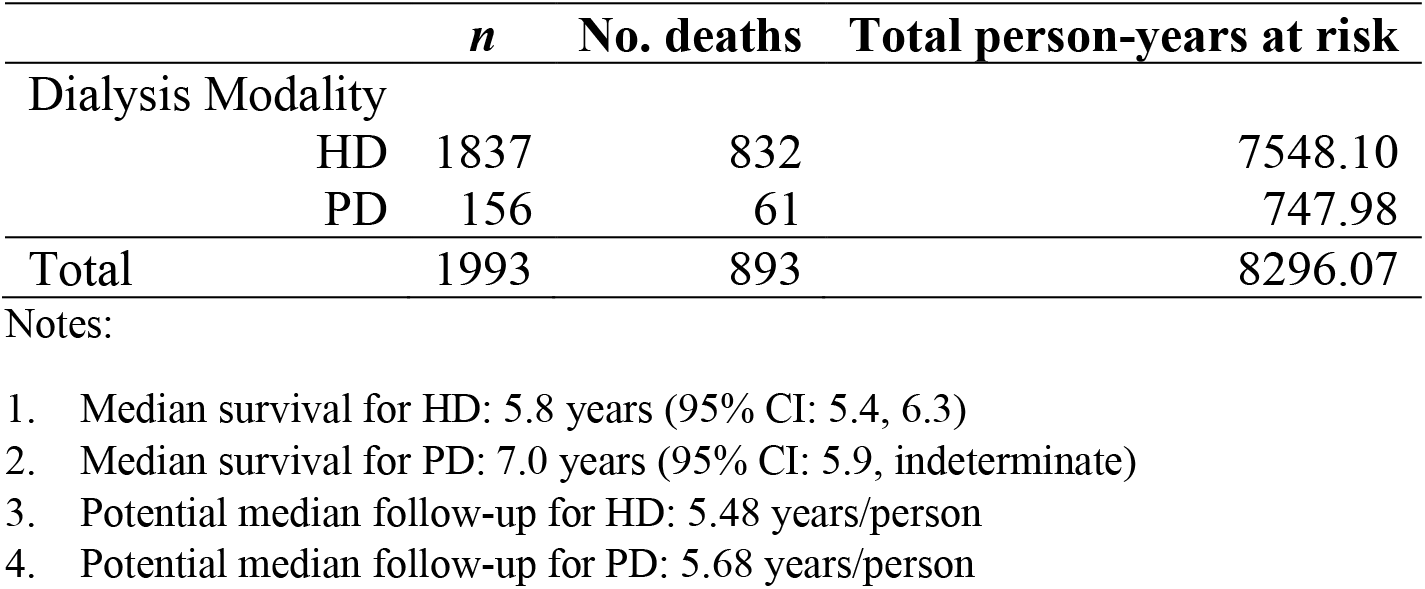
Number of deaths and total person-years at risk (*n* = 1993).

The majority of the patients were started on HD. The distributions of demographic characteristics and laboratory values among patients who were started on PD were similar to those of patients who were started on HD as shown in Table 4. All Cohen’s *d* values were less than |0.5|. Without adjusting for any confounding variables, there was evidence of a survival benefit of starting dialysis with PD versus with HD. Figure 2 shows the unadjusted Kaplan-Meier survival curves comparing PD versus with HD for both all cases and complete cases. Table 5 shows that the effect of dialysis modality was not significant at the 5 percent level of significance after controlling for confounders. The rate of death among those who started with PD could be as much as 39 percent lower or 5 percent higher than that of those who started with HD with 95 percent confidence.

**Table 4.** Comparison of patient characteristics between HD and PD groups (*n* = 1993).

|  | **HD ($n = 1837$)** | **PD ($n = 156$)** | ***t*-stat. (*df*)<sup>a</sup>** | ***p*-value<sup>a</sup>** | **Cohen's *d*** |
| --- | --- | --- | --- | --- | --- |
|  | **Mean (SD)** |  |  |  |  |
| Age (years) | 54.27 (13.34) | 48.15 (14.13) | 5.22 (179.29) | < 0.001 | 0.46 |
| BMI (kg/m<sup>2</sup>) | 24.81 (4.69) | 24.12 (4.41) | 1.87 (186.08) | 0.064 | 0.15 |
| Total Cholesterol (mmol/L) | 4.39 (1.14) | 4.60 (1.07) | -2.35 (185.97) | 0.020 | -0.19 |
| Serum Creatinine (μm/L) | 811.37 (291.73) | 823.40 (249.96) | -0.57 (192.71) | 0.570 | -0.04 |
| Serum Albumin (g/L) | 37.32 (6.51) | 35.95 (6.32) | 2.59 (184.12) | 0.010 | 0.21 |
| Serum Calcium (mmol/L) | 2.18 (0.24) | 2.16 (0.21) | 0.97 (193.01) | 0.334 | 0.07 |
| Serum Phosphate (mmol/L) | 1.79 (0.55) | 1.67 (0.46) | 3.23 (194.23) | 0.001 | 0.23 |
| Serum ALP (U/L) | 113.46 (80.42) | 110.14 (68.46) | 0.57 (193.22) | 0.568 | 0.04 |
| Serum Potassium (mmol/L) | 4.52 (0.85) | 4.11 (0.66) | 7.31 (202.22) | < 0.001 | 0.49 |
| Hb (g/dL) | 9.81 (1.68) | 10.31 (1.56) | -3.79 (186.91) | < 0.001 | -0.30 |
|  | ***n* (%)** |  | **χ<sup>2</sup>-stat. (*df*)<sup>b</sup>** | ***p*-value<sup>b</sup>** |  |
| Sex |  |  | 5.51 (1) | 0.019 |  |
| Male | 1056 (57) | 74 (47) |  |  |  |
| Female | 781 (43) | 82 (53) |  |  |  |
| DM Comorbidity |  |  | 0.82 (1) | 0.364 |  |
| Yes | 959 (52) | 75 (48) |  |  |  |
| No | 878 (48) | 81 (52) |  |  |  |
| CVD Comorbidity |  |  | 2.09 (1) | 0.148 |  |
| Yes | 135 (7) | 17 (11) |  |  |  |
| No | 1702 (93) | 139 (89) |  |  |  |

a Welch's two-sample *t* test; b Chi square test of independence

**Table 5.** Assessment of the effect of PD versus that of HD using WCR (*n* = 1993).

|  |  | *b* (SE) | AHR (95% CI) | *z*-stat. | *p*-value |
| --- | --- | --- | --- | --- | --- |
| **Model 1** | PD | -0.340 (0.131) | 0.712 (0.551, 0.920) | -2.60 | 0.009 |
| **Model 2** | PD | -0.226 (0.139) | 0.798 (0.608, 1.048) | -1.62 | 0.104 |

Notes:
1. WCR was used to account for nonproportional hazards. 2. Model 1 was unadjusted. 3. Model 2 was adjusted for age, sex, DM comorbidity, CVD comorbidity, BMI, serum creatinine, serum albumin, serum calcium, serum phosphate, serum potassium, serum ALP, total cholesterol, and Hb.

**Figure 2.**
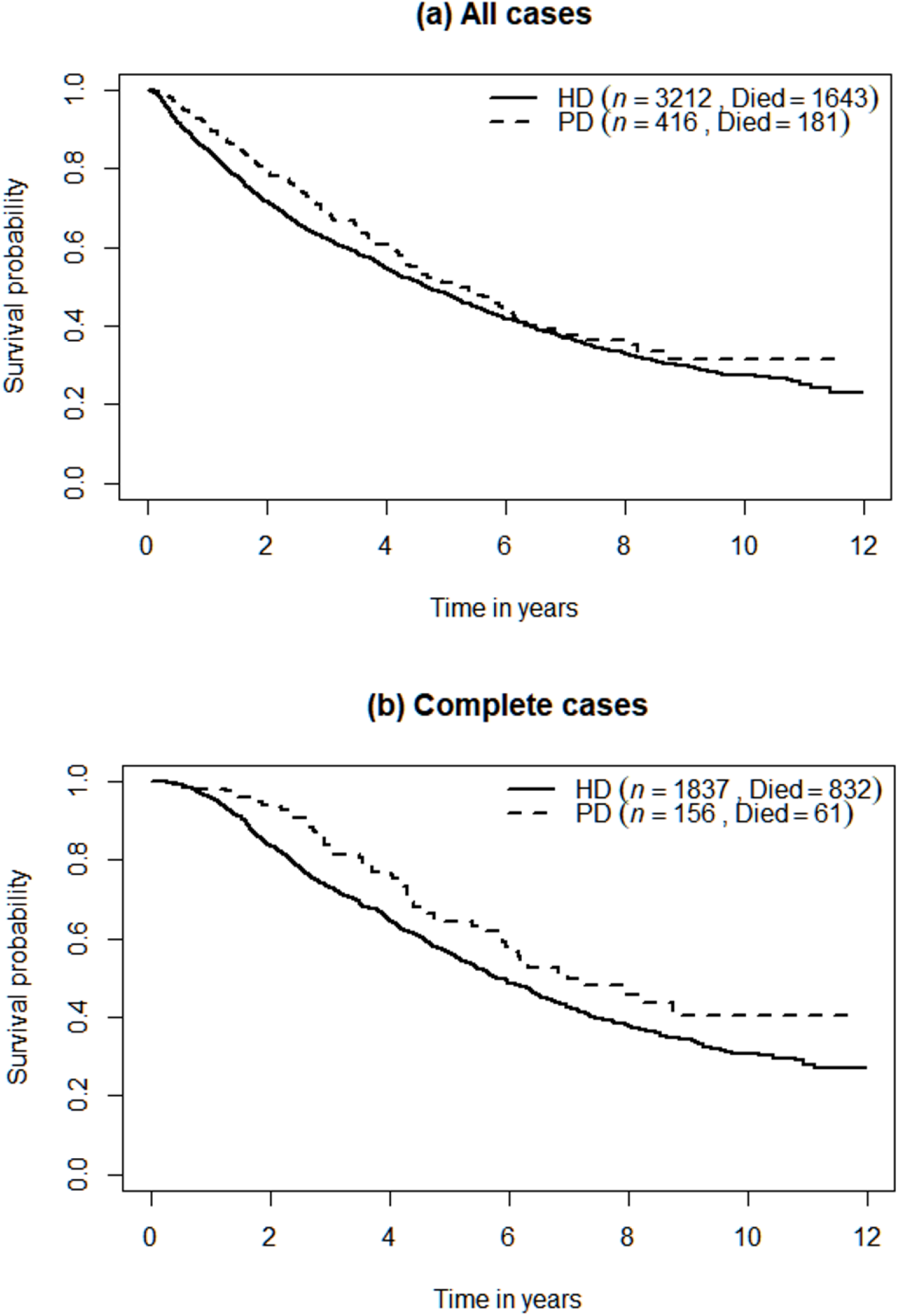
Unadjusted Kaplan-Meier survival curves comparing PD versus with HD for both all cases and complete cases.

As shown in Table 6, it appears that older age, male sex, the presence of DM, higher serum phosphate, and higher serum ALP were significantly associated with a higher death rate. On the other hand, higher serum creatinine, higher serum albumin, higher serum potassium, higher Hb, and higher BMI were all significantly associated with a lower death rate.

**Table 6.** The most parsimonious WCR model (*n* = 1993).

| **Model 3** | *b* (SE) | AHR (95% CI) | *z*-stat. | *p*-value |
| --- | --- | --- | --- | --- |
| PD <sub>vs</sub> HD | -0.232 (0.139) | 0.793 (0.604, 1.043) | -1.66 | 0.097 |
| Age | 0.488 (0.041) | 1.630 (1.504, 1.766) | 11.96 | < 0.001 |
| Male <sub>vs</sub> Female | 0.210 (0.075) | 1.233 (1.064, 1.429) | 2.79 | 0.005 |
| DM comorbidity <sub>vs</sub> No DM | 0.241 (0.075) | 1.272 (1.099, 1.472) | 3.22 | 0.001 |
| CVD comorbidity <sub>vs</sub> No CVD | 0.239 (0.122) | 1.269 (1.000, 1.612) | 1.96 | 0.050 |
| BMI | -0.105 (0.041) | 0.900 (0.831, 0.975) | -2.57 | 0.010 |
| Serum creatinine | -0.161 (0.047) | 0.851 (0.776, 0.933) | -3.43 | < 0.001 |
| Serum albumin | -0.240 (0.032) | 0.787 (0.739, 0.838) | -7.52 | < 0.001 |
| Serum phosphate | 0.110 (0.035) | 1.117 (1.043, 1.195) | 3.19 | 0.001 |
| Serum ALP | 0.078 (0.033) | 1.081 (1.013, 1.153) | 2.35 | 0.019 |
| Serum potassium | -0.089 (0.041) | 0.915 (0.844, 0.991) | -2.17 | 0.030 |
| Hb | -0.252 (0.039) | 0.777 (0.721, 0.838) | -6.54 | < 0.001 |

Notes:
1. All continuous covariates were standardized. Therefore, each regression coefficient represents the change in log relative hazard per SD change in the covariate. 2. There were no significant interactions between dialysis modality, age, DM comorbidity, and CVD comorbidity. 3. All continuous covariates were found to be approximately linear against log relative hazard. 4. There were no influential outliers identified. 5. The overall goodness-of-fit was acceptable.

## Discussion

This is the largest contemporary study to date on survival in treated ESRD patients in Sabah who began treatment with either HD or PD. The study used the MDTR database, the largest registered database of treated ESRD patients in Malaysia, to compare the survival of ESRD patients who started on HD with that of those who started on PD. Moreover, the effects of other predictors on survival in this population were also studied.

The study demonstrated that initial dialysis modality did not significantly impact all-cause mortality of dialysis patients in Sabah after controlling for selected confounders. This finding is in agreement with what was found in the whole Malaysia cohort [12] and is corroborated by large observational studies [4,7,17,18]. Conflicting results are numerous, however. The inconsistencies may be explained by methodologic differences, such as the type of statistical models used, degree of adjustment, selection of subjects, and length of follow up. However, it could also be due to the absence of a true difference between both modalities [5,19]. Given that PD as an initial dialysis modality was found to be more cost-effective than HD [20], the finding from this study, if confirmed, further strengthens the argument for the increased use of PD in the local setting.

Unsurprisingly, it was found that older age and the presence of DM were significantly associated with poorer survival. The death rate was 27% (95% CI: 10%, 47%) higher in diabetic ESRD patients and increased by 63% (95% CI: 50%, 77%) for every SD, approximately 14 years, increase in age. This is consistent with the conclusions from most studies [21]. In fact, Locatelli et al. [22] claimed that age is the most important demographic factor associated with increased mortality, together with the increasing incidence of diabetic patients with ESRD. Older patients are frailer and more susceptible to concomitant medical conditions. Moreover, DM is a predictor of CVD which accounts for more than 50% of mortality in these patients, leading to a mortality rate approximately 30 times higher than that in the general population [23].

Another expected finding is that higher serum albumin was significantly associated with lower all-cause mortality in these patients. The death rate decreased by 21% for every SD, approximately 6.5 g/L, increase in serum albumin. A decrease in the death rate of between 16% and 26% is consistent with the data at the 95% confidence level. Low serum albumin is a predictor of mortality regardless of modality of RRT [3]. Serum albumin is a marker of nutritional status and low serum albumin is one of the criteria for protein energy wasting (PEW) which is often associated with mortality [24]. This finding suggests that hypoalbuminemia should be avoided to reduce the risk of death in these patients.

The effects of other variables on the survival of dialysis patients are unclear. First, this study found that the rate of death among males was 23% higher than that among females; and could be as little as 6% higher to as much as 43% higher with 95% confidence. This finding is in agreement with what was reported by Wang et al. [25]. However, many studies have reported that sex has no significant effect on the survival of dialysis patients [17,22,26,27]. The result does not necessarily imply anything important as it may just reflect the fact that females have greater survival than males in the general population.

Second, it was found that BMI was an independent predictor of death in dialysis patients. The rate of death decreased by 10% for every SD, approximately 5 kg/m^2^, increase in BMI. A decrease in rate of death of between 2% and 17% is consistent with the data at the 95% confidence level. This finding is consistent with what was reported by Wu et al. [21]. Contrary to the finding in the general population, in which high BMI is associated with high mortality, this phenomenon of “reverse epidemiology” in which low BMI is associated with high mortality was discussed in length by Kalantar-Zadeh et al. [28]. Possible explanations include a more stable hemodynamic status, alterations in circulating cytokines, unique neurohormonal constellations, endotoxin-lipoprotein interaction, reverse causation, survival bias, time discrepancies among competitive risk factors, and malnutrition-inflammation complex syndrome [28]. Weight-gain-related interventional studies in dialysis patients are needed to confirm this finding.

Third, this study shows that serum creatinine was inversely correlated with mortality risk. The rate of death decreased by 15% for every SD, approximately 289 µm/L, increase in serum creatinine. A decrease in the rate of death of between 7% and 22% is consistent with the data at 95% confidence level. This paradox is consistent with the results of past studies [24,29]. The deterioration in muscle mass and nutritional status in ESRD patients may confound the distribution of the creatinine value in this population, with the healthiest patients having the greatest creatinine values and the sickest patients with the latest referral for dialysis having the lowest values [30]. The implication of this finding is that serum creatinine is not a reliable surrogate for residual renal function in new ESRD patients, but it can be used as a predictor of subsequent mortality in this population.

Fourth, this study reveals that out of the three biomarkers of chronic kidney disease-mineral and bone disorder (CKD-MBD) studied, only serum phosphate and ALP were significantly associated with mortality in dialysis patients. The death rate increased by 12% (95% CI: 4%, 20%) and 8% (95% CI: 1%, 15%) for every SD increase in serum phosphate and ALP, respectively. However, serum calcium was found to be not significantly related to mortality. The result is consistent with what was reported by Owaki et al. [31], but there are other studies that yielded differing results [32,33]. The discrepancies may be attributed to the different measures used by different studies. Some studies used aggregate measures of laboratory values over time whereas others used single measures at baseline [32]. This result implies that bone mineral parameters especially phosphate and ALP need to be kept low to reduce the death rate among dialysis patients.

Fifth, this study shows that serum potassium was significantly associated with mortality in dialysis patients. The death rate decreased by as little as 1% to as much as 16% for every SD, approximately 0.84 mmol/L, increase in serum potassium with 95% confidence. This seemingly linear relationship is only true for the observed data. A more plausible relationship is the U-shaped relationship. Eriguchi et al. [34] reported that very low and very high serum potassium levels were associated with higher all-cause mortality in incident PD patients, which can be partly explained by the impact of potassium on ventricular arrhythmogenicity. However, a study by Oliveira et al. [35] in HD patients found that serum potassium was not a significant predictor of mortality. The findings from this study suggest that hypokalemia should be avoided in dialysis patients.

Last, it was found that Hb was an independent predictor of mortality in dialysis patients. The death rate decreased by as little as 16% to as much as 28% for every SD, approximately 1.67 g/dL, increase in Hb with 95% confidence. Abe et al. [36] reported that Hb was not a significant predictor of mortality in PD patients. In another study among HD patients, it was shown that a low Hb concentration was a robust hazard factor, whereas a high Hb concentration did not increase the risk. The mortality risk rapidly increased when the Hb concentration was below 10.0 g/dL [37]. Low Hb leads to anemia, a common complication of CKD due to reduced erythropoietin production and iron absorption by the kidney. This result implies that low Hb concentration should be prevented to reduce the mortality risk in this population.

One important strength of the current study is the way in which the censored survival data were analyzed. The Cox PH regression model is the most popular method in the analysis of such data. However, the assumption of proportional hazards is often violated resulting in overestimation or underestimation of the hazard ratios. To cope with the nonproportional hazards, WCR was used in this study. The inclusion of parameters for time-dependent effects is a viable alternative but it usually results in a more complex model. Additionally, there is little medical interest in the time-dependence of a covariate effect [15]. Stratification based on a prognostic factor exhibiting nonproportional hazards is not an attractive option as it precludes estimation of its effect. Furthermore, stratification is not viable when many prognostic factors exhibit nonproportional hazards. In contrast, WCR can simplify analysis and interpretation. Via weighted estimation, a direct comparison of the magnitude of effects from proportional and nonproportional covariates of interest can be performed [15].

The study has two important limitations. First, the regression analyses were based on complete cases. As many as 45 percent of patients have been excluded due to missing data for at least one of the variables considered for regression analyses. Although the characteristics of the complete cases were representative of all cases, as shown in Table 1 and Table 2, the sample size was reduced substantially, resulting in a loss of efficiency and increased standard errors. Multiple imputation is a viable alternative to deal with missing data, but it is best viewed in the context of sensitivity analysis [38]. Future studies should employ both methods and compare the results.

Second, the laboratory values measured were not baseline values. Instead, they were averages of repeated measurements over time measured after dialysis initiation. The aggregation of repeated measures into summary statistics such as the mean, median or extreme values common in studies whose data sources are existing health records [39]. The summarization of longitudinal covariates may be a missed analytic opportunity. The incorporation of these variables as time-varying covariates in the model may be more appropriate. On the other hand, Sweeting et al. [40] demonstrated that simpler summarization-based approaches were just as effective as more sophisticated methods and modestly better than the baseline-only model. Future studies should consider models with time-varying covariates when dealing with longitudinal variables that fluctuate strongly over time. Otherwise, simple summary measures should suffice.

The patients studied are representative of all adult ESRD patients who received dialysis treatment in the dialysis centers in Sabah that participated in the MDTR. The generalization of the results to all adult ESRD patients receiving dialysis treatment in Sabah is not unreasonable given that there is no reason to suspect any dialysis centers not reporting to the MDTR. Most of the patients received their treatment in the government settings, all of which participated in the MDTR. The few private or non-profit organization (NGO) centers that exist in Sabah can be considered compliant to data submission to the MDTR given the Private Healthcare Facilities Act 1998 (AKTA 586) that states that all dialysis health facilities are required to submit data to the MDTR. Any significant associations found in this study do not at all imply causation. The significant predictors of all-cause mortality identified may or may not be the causal factors.

In conclusion, it was found that dialysis modality was not significantly related to all-cause mortality after controlling for confounders. The most parsimonious regression model identified age, sex, DM comorbidity, BMI, serum creatinine, serum albumin, serum phosphate, serum ALP, serum potassium, and Hb as statistically significant predictors of survival.

## Data Availability

The datasets generated during and/or analysed during the current study are available from the corresponding author on reasonable request.

## Ethical disclosures

This study has been approved by Medical Research & Ethics Committee (MREC) and the Human Research Ethics Committee USM (HREC). The registration numbers were NMRR-19-1120-48161 (IIR) and USM/JEPeM/19120840, respectively.

## Funding

Self-funded.

## Conflicts of interest/Competing interests

None declared.

## Acknowledgments

The authors would like to thank the Director General of Health of Malaysia for permission to publish the results of this study. The authors would also like to thank the National Renal Registry (NRR) for the data provided.

## Notes

### Competing Interest Statement

The authors have declared no competing interest.

## References

1. National Institutes of Health, National Institute of Diabetes and Digestive and Kidney Diseases (2017) United States Renal Data System. 2017 USRDS annual data report: Epidemiology of kidney disease in the United States. https://www.usrds.org/2018/view/v2_11.aspx. Accessed 9 May 2019 2019

2. Vale L, Cody JD, Wallace SA, Daly C, Campbell MK, Grant AM, Khan I, MacLeod AM (2004) Continuous ambulatory peritoneal dialysis (CAPD) versus hospital or home haemodialysis for end-stage renal disease in adults. Cochrane Database Syst Rev (4)

3. Singh T, Astor BC, Waheed S (2019) END-STAGE RENAL DISEASE PATIENTS WITH LOW SERUM ALBUMIN: IS PERITONEAL DIALYSIS AN OPTION? Perit Dial Int 39 (6):562–567. doi:10.3747/pdi.2018.00204

4. Zhou H, Sim JJ, Bhandari SK, Shaw SF, Shi J, Rasgon SA, Kovesdy CP, Kalantar-Zadeh K, Kanter MH, Jacobsen SJ (2019) Early Mortality Among Peritoneal Dialysis and Hemodialysis Patients Who Transitioned With an Optimal Outpatient Start. Kidney Int Rep 4 (2):275–284. doi:10.1016/j.ekir.2018.10.008

5. Korevaar JC, Feith GW, Dekker FW, van Manen JG, Boeschoten EW, Bossuyt PM, Krediet RT (2003) Effect of starting with hemodialysis compared with peritoneal dialysis in patients new on dialysis treatment: a randomized controlled trial. Kidney Int 64 (6):2222–2228. doi:10.1046/j.1523-1755.2003.00321.x

6. Kumar VA, Sidell MA, Jones JP, Vonesh EF (2014) Survival of propensity matched incident peritoneal and hemodialysis patients in a United States health care system. Kidney Int 86 (5):1016–1022. doi:10.1038/ki.2014.224

7. Wong B, Ravani P, Oliver MJ, Holroyd-Leduc J, Venturato L, Garg AX, Quinn RR (2018) Comparison of Patient Survival Between Hemodialysis and Peritoneal Dialysis Among Patients Eligible for Both Modalities. Am J Kidney Dis 71 (3):344–351. doi:10.1053/j.ajkd.2017.08.028

8. Lee MJ, Kwon YE, Park KS, Kee YK, Yoon CY, Han IM, Han SG, Oh HJ, Park JT, Han SH, Yoo TH, Kim YL, Kim YS, Yang CW, Kim NH, Kang SW (2016) Glycemic Control Modifies Difference in Mortality Risk Between Hemodialysis and Peritoneal Dialysis in Incident Dialysis Patients With Diabetes: Results From a Nationwide Prospective Cohort in Korea. Medicine 95 (11):e3118. doi:10.1097/md.0000000000003118

9. Wang IK, Lin CL, Yen TH, Lin SY, Sung FC (2018) Comparison of survival between hemodialysis and peritoneal dialysis patients with end-stage renal disease in the era of icodextrin treatment. Eur J Intern Med 50:69–74. doi:10.1016/j.ejim.2017.11.017

10. Lee SW, Lee NR, Son SK, Kim J, Sul AR, Kim Y, Park JT, Lee JP, Ryu DR (2019) Comparative study of peritoneal dialysis versus hemodialysis on the clinical outcomes in Korea: a population-based approach. Sci Rep 9 (1):5905. doi:10.1038/s41598-019-42508-z

11. Yang F, Khin LW, Lau T, Chua HR, Vathsala A, Lee E, Luo N (2015) Hemodialysis versus Peritoneal Dialysis: A Comparison of Survival Outcomes in South-East Asian Patients with End-Stage Renal Disease. PloS one 10 (10):e0140195. doi:10.1371/journal.pone.0140195

12. National Renal Registry (2018) 24th Report of the Malaysian Dialysis and Transplant Registry 2016. https://www.msn.org.my/nrr/mdtr2016.jsp. Accessed 9 May 2019 2019

13. Stack AG (2002) Determinants of modality selection among incident US dialysis patients: results from a national study. J Am Soc Nephrol 13 (5):1279–1287

14. Moore DF (2016) Applied survival analysis using R. Springer, California

15. Dunkler D, Ploner M, Schemper M, Heinze G (2018) Weighted Cox regression using the R package coxphw. J Stat Softw 84 (1):1–26

16. Schemper M, Wakounig S, Heinze G (2009) The estimation of average hazard ratios by weighted Cox regression. Stat Med 28 (19):2473–2489

17. Mircescu G, Stefan G, Garneata L, Mititiuc I, Siriopol D, Covic A (2014) Outcomes of dialytic modalities in a large incident registry cohort from Eastern Europe: the Romanian Renal Registry. Int Urol Nephrol 46 (2):443–451. doi:10.1007/s11255-013-0571-3

18. Haapio M, Helve J, Kyllonen L, Gronhagen-Riska C, Finne P (2013) Modality of chronic renal replacement therapy and survival--a complete cohort from Finland, 2000-2009. Nephrol Dial Transplant 28 (12):3072–3081. doi:10.1093/ndt/gft326

19. Vonesh EF, Snyder JJ, Foley RN, Collins AJ (2004) The differential impact of risk factors on mortality in hemodialysis and peritoneal dialysis. Kidney Int 66 (6):2389–2401. doi:10.1111/j.1523-1755.2004.66028.x

20. Surendra NK, Abdul Manaf MR, Hooi LS, Bavanandan S, Mohamad Nor FS, Firdaus Khan SS, Meng OL, Abdul Gafor AH (2019) Cost utility analysis of end stage renal disease treatment in Ministry of Health dialysis centres, Malaysia: Hemodialysis versus continuous ambulatory peritoneal dialysis. PloS one 14 (10):e0218422. doi:10.1371/journal.pone.0218422

21. Wu B, Wang M, Gan L, Zhao H (2014) Comparison of patient survival between hemodialysis and peritoneal dialysis in a single Chinese center. Int Urol Nephrol 46 (12):2403–2407. doi:10.1007/s11255-014-0819-6

22. Locatelli F, Marcelli D, Conte F, Del Vecchio L, Limido A, Malberti F, Spotti D, Sforzini S, for the Registro Lombardo Dialisi e T (2000) Patient selection affects end-stage renal disease outcome comparisons. Kidney Int 57:S94–S99. doi:10.1046/j.1523-1755.2000.07416.x

23. Levey AS, Beto JA, Coronado BE, Eknoyan G, Foley RN, Kasiske BL, Klag MJ, Mailloux LU, Manske CL, Meyer KB, Parfrey PS, Pfeffer MA, Wenger NK, Wilson PW, Wright JT, Jr. (1998) Controlling the epidemic of cardiovascular disease in chronic renal disease: what do we know? What do we need to learn? Where do we go from here? National Kidney Foundation Task Force on Cardiovascular Disease. Am J Kidney Dis 32 (5):853–906. doi:10.1016/s0272-6386(98)70145-3

24. Kanda E, Kato A, Masakane I, Kanno Y (2019) A new nutritional risk index for predicting mortality in hemodialysis patients: Nationwide cohort study. PloS one 14 (3):e0214524. doi:10.1371/journal.pone.0214524

25. Wang IK, Kung PT, Kuo WY, Tsai WC, Chang YC, Liang CC, Chang CT, Yeh HC, Wang SM, Chuang FR, Wang KY, Lin CY, Huang CC (2013) Impact of dialysis modality on the survival of end-stage renal disease patients with or without cardiovascular disease. J Nephrol 26 (2):331–341. doi:10.5301/jn.5000149

26. Sens F, Schott-Pethelaz AM, Labeeuw M, Colin C, Villar E (2011) Survival advantage of hemodialysis relative to peritoneal dialysis in patients with end-stage renal disease and congestive heart failure. Kidney Int 80 (9):970–977. doi:10.1038/ki.2011.233

27. Waldum-Grevbo B, Leivestad T, Reisaeter AV, Os I (2015) Impact of initial dialysis modality on mortality: a propensity-matched study. BMC Nephrol 16:179. doi:10.1186/s12882-015-0175-5

28. Kalantar-Zadeh K, Abbott KC, Salahudeen AK, Kilpatrick RD, Horwich TB (2005) Survival advantages of obesity in dialysis patients. Am J Clin Nutr 81 (3):543–554. doi:10.1093/ajcn/81.3.543

29. Xue JL, Everson SE, Constantini EG, Ebben JP, Chen S-C, Agodoa LY, Collins AJ (2002) Peritoneal and hemodialysis: II. Mortality risk associated with initial patient characteristics. Kidney Int 61 (2):741–746

30. Fink JC, Burdick RA, Kurth SJ, Blahut SA, Armistead NC, Turner MS, Shickle LM, Light PD (1999) Significance of serum creatinine values in new end-stage renal disease patients. Am J Kidney Dis 34 (4):694–701. doi:10.1016/s0272-6386(99)70395-1

31. Owaki A, Inaguma D, Tanaka A, Shinjo H, Inaba S, Kurata K (2017) Evaluation of the Relationship between the Serum Alkaline Phosphatase Level at Dialysis Initiation and All-Cause Mortality: A Multicenter, Prospective Study. Nephron Extra 7 (3):78–88. doi:10.1159/000481409

32. Melamed ML, Eustace JA, Plantinga L, Jaar BG, Fink NE, Coresh J, Klag MJ, Powe NR (2006) Changes in serum calcium, phosphate, and PTH and the risk of death in incident dialysis patients: a longitudinal study. Kidney Int 70 (2):351–357. doi:10.1038/sj.ki.5001542

33. Lee JE, Lim JH, Jang HM, Kim YS, Kang SW, Yang CW, Kim NH, Kwon E, Kim HJ, Park JM, Jung HY, Choi JY, Park SH, Kim CD, Cho JH, Kim YL (2017) Low serum phosphate as an independent predictor of increased infection-related mortality in dialysis patients: A prospective multicenter cohort study. PloS one 12 (10):e0185853. doi:10.1371/journal.pone.0185853

34. Eriguchi R, Obi Y, Soohoo M, Rhee CM, Kovesdy CP, Kalantar-Zadeh K, Streja E (2019) Racial and Ethnic Differences in Mortality Associated with Serum Potassium in Incident Peritoneal Dialysis Patients. Am J Nephrol:1–9. doi:10.1159/000502998

35. Oliveira TS, Valente AT, Caetano CG, Garagarza CA (2017) Nutritional parameters as mortality predictors in haemodialysis: Differences between genders. J Ren Care 43 (2):83–91. doi:10.1111/jorc.12201

36. Abe M, Hamano T, Hoshino J, Wada A, Nakai S, Hanafusa N, Masakane I, Nitta K, Nakamoto H (2019) Predictors of outcomes in patients on peritoneal dialysis: A 2-year nationwide cohort study. Sci Rep 9 (1):3967. doi:10.1038/s41598-019-40692-6

37. Jin DC (2019) Analysis of mortality risk from Korean hemodialysis registry data 2017. Kidney Res Clin Pract 38 (2):169–175. doi:10.23876/j.krcp.19.011

38. Vandenbroucke JP, von Elm E, Altman DG, Gotzsche PC, Mulrow CD, Pocock SJ, Poole C, Schlesselman JJ, Egger M (2014) Strengthening the Reporting of Observational Studies in Epidemiology (STROBE): explanation and elaboration. Int J Surg 12 (12):1500–1524. doi:10.1016/j.ijsu.2014.07.014

39. Goldstein BA, Pomann GM, Winkelmayer WC, Pencina MJ (2017) A comparison of risk prediction methods using repeated observations: an application to electronic health records for hemodialysis. Stat Med 36 (17):2750–2763. doi:10.1002/sim.7308

40. Sweeting MJ, Barrett JK, Thompson SG, Wood AM (2017) The use of repeated blood pressure measures for cardiovascular risk prediction: a comparison of statistical models in the ARIC study. Stat Med 36 (28):4514–4528

